# Preventable maternal deaths in England and Wales, 2013-2023: a systematic case series of coroners’ reports

**DOI:** 10.1101/2024.07.09.24310137

**Authors:** Jessy Jindal, David Launer, Georgia C. Richards, Francesco Dernie

## Abstract

**Objective:** Coroners in England and Wales have a duty to write Prevention of Future Deaths (PFDs) reports when they believe that action should be taken to prevent similar deaths. We aimed to characterise learnings from reports involving maternal deaths.

**Design:** Systematic case series

**Setting:** England and Wales

**Population or Sample:** Database of all coroners’ PFDs published between July 2013 and 1 August 2023. There were 4435 reports at the time of data collection.

**Methods:** A reproducible computer code developed from the Preventable Deaths Tracker (https://preventabledeathstracker.net/) was used to download all published PFDs from the Judiciary website. Reports were searched for keywords related to maternal deaths. Case information was extracted into pre-specified domains and compared to other data on maternal deaths.

**Main Outcome Measures:** Case demographics, causes of deaths, risk factors, coroner concerns and organisational responses.

**Results:** Twenty nine reports involved a maternal death. The median age at death was 33.5 years (IQR 29-36 years) and 76% of deaths occurred in hospitals. The most common cause of death was haemorrhage. Coroners frequently voiced concerns around failure to provide appropriate treatment (57%), and failure of timely escalation (38%). Only 38% of PFDs had published responses. When organisations did respond to the coroner, 80% reported that they implemented changes, including publishing new local policies, increasing training, or committing to increased staffing.

**Conclusions:** PFDs highlight gaps in obstetric care which, if appropriately addressed, and regularly and routinely monitored, could prevent similar deaths.

**Funding:** None

## Introduction

Global maternal deaths remain high and have stalled or worsened in 150 countries since 2000^1^. In 2020, the global maternal mortality rate was 223 deaths per 100,000 live births, which corresponds approximately to one maternal death every 2 minutes^1^. In the United Kingdom (UK), between 2019 and 2021, the Mothers and Babies Reducing Risk through Audits and Confidential Enquiries (MBRRACE) programme reported 241 women died during or within 42 days of their pregnancy, a rate of 11.7 deaths per 100,000 maternities^2^. However, maternal deaths have not reduced in the past decade in the UK, and there is inequality in mortality rates for women from areas of economic deprivation and ethnic minorities^2^. As a result, the UK government commitment to ending maternity-related preventable deaths of both mothers and their children by 2030^3^.

Coroners in England and Wales have a duty to report when they believe that action should be taken to prevent future deaths^4^. These reports are called Prevention of Future Deaths reports or PFDs^4,5^, which are sent to addressees who must respond to the coroner within 56 days. PFDs are listed by the National Health Service (NHS) Patient Safety Strategy as an official source of data^6^, but it is unclear how they are used, if they are monitored, and what action is taken to prevent similar fatalities in the NHS. A detailed analysis of PFDs implicating maternal deaths could highlight factors contributing to maternal mortality and continuing inequalities in care. This would help inform clinical staff managing pregnanies and public health policy by highlighting points for improvement in care.

The aim of our study was to systematically determine the number of maternal deaths reported by coroners in PFDs and to characterise these deaths in terms of demographics, risk factors and causes of deaths. In addition, we aimed to explore concerns raised by coroners in these reports and understand what actions were reported by organisations in their responses to the coroner.

## METHODS

The systematic case series was designed and a study protocol was developed and preregistered on an open repository^7^.

### Data collection, screening and eligibility

We used a reproducible, openly available code written by FD to download all portable document format (pdf) documents from the judiciary website published from inception (July 2013) to 1 August 2023. The code was reproduced from the Preventable Deaths Tracker (https://preventabledeathstracker.net/) and is available here: https://github.com/francescodernie/coroner_PFDs.

We used the OCCRP Aleph tool^8^ to create a repository of the PFDs to process the reports, allowing them to be machine-readable. We then conducted keyword searches using the following terms: ‘pregnancy’, ‘pregnant’, ‘maternal’, ‘post-partum’, ‘partum’, ‘natal’’, ‘perinatal’, ‘antenatal’, ‘obstetrics’, ‘gestation’, ‘parturition, ‘birth’, and the positive control word ‘coroner’ (to identify documents that could not be read automatically).

Cases were included when a maternal death occurred, using the World Health Organisation (WHO) definition^9^: deaths of a female due to any causes related to or aggravated by pregnancy or its management, both during pregnancy, childbirth or within 42 days of its resolution. We also included late maternal deaths defined as “the death of a woman from direct or indirect obstetric causes, more than 42 days but less than one year after termination of pregnancy”.^9^ Deaths were excluded if the incidental causes of death were unrelated to the pregnancy (e.g. a road traffic accident) as per the WHO definition.^9^

### Data extraction

Data was extracted from the included PFDs in line with established methodology that has been used in other case series of PFDs^10–12^. Demographics, causes of deaths, pregnancy outcomes and risk factors, coroner concerns and organisational responses were extracted manually by two authors (JJ, DL) and reviewed by FD. Risk factors extracted were based on existing literature^13^, including gestational diabetes, hypertensive disease, obesity, cardiac disease, haematological disorders, psychiatric illness, epilepsy, aged <17 years or >35 years, recreational drug use, complications from previous pregnancy, multiple pregnancies, hepatic disease, and congenital anomalies of the child.

The numbers of maternal deaths (up to 1 year from birth) from MBRRACE-UK^2^ were also extracted from each MBRRACE-UK report between 2014-2021, to compare to the number of PFDs in the same time period. Rolling averages over 3 year periods were taken to match the presentation of the MBRRACE-UK data.

### Data analysis

The number of maternal deaths reported in coroners’ PFDs and their rates as a proportion of the maternal deaths reported in MBRRACE-UK were calculated over time. Medians and interquartile ranges (IQRs) were calculated for continuous variables (e.g age) and frequencies were reported for categorical variables (e.g sex, location of death, and coroner jurisdiction area).

We calculated the years of life lost (YLL)^14^ for each case (where age was reported) by extracting their remaining life expectancy from the ONS cohort life tables^15^. The cause of death determined by the coroner in each case was assigned to categories stipulated in the 2019-21 MBRRACE-UK report^2^. Two investigators (JJ, DL) also assigned the International Statistical Classification of Diseases and Related Health Problems 11th Revision (ICD-11) numeric codes for the causes of death to each PFD^16^.

Directed content analysis^17^ was used to collate and evaluate concerns raised by coroner and classify them. Concerns were classified by one author (JJ) and ambiguities clarified with other authors (DL, FD).

To calculate response rates to PFDs, we used the 56-day legal requirement to classify responses as “early or on time” (on or before the due date), “late” (after due date), or “overdue” (response was not available on the Judiciary website as of the time of extraction). We calculated the average response rate and frequency for recipients. The content of responses were classified by the type of change reported if applicable.

### Missing data

Coroners have a duty to write PFDs^4,5^, but this is not mandatory or enforced. Thus, data is constrained by the working practices of coroners who may vary in their thresholds for writing a PFD report. Furthermore, PFDs must be sent to the Chief Coroner’s Office who will assess the report before publication. In some instances, reports may go unpublished as they have not been sent to the Chief Coroner’s Office, get lost in the email inboxes or, in very rare cases the Chief Coroner will choose not publish the report at the request of family members or if the report poses a risk to the public^18^. We can therefore only analyse the publicly-available PFDs.

### Patient involvement

No patients were directly involved in this study, however our research team constantly engages with bereaved families and friends who have lost a loved one through our platform the Preventable Deaths Tracker (https://preventabledeathstracker.net/). This engagement has guided the development of our platform and helped identify new research projects on specific areas of death prevention.

### Software and data sharing

We used R (version 4.1.1) to create the openly available code to download and screen the pdf documents^19^. The OCCRP Aleph tool, an openly available platform was used for document storage and investigation management, as well as conducting the keyword searches^8^. Microsoft Excel was used for data extraction and analysis. Figures were created using Datawrapper^20^.

## RESULTS

There were 29 coroners’ PFDs involving maternal deaths between July 2013 and August 2023 in England and Wales (0.7% of all available PFDs, n=4435). When compared to data from MBRRACE-UK, PFDs represented only a small fraction of total maternal deaths (Table 1). Direct comparison was difficult due to MBRRACE data including all UK deaths, whereas PFDs only cover England and Wales.

**Table 1.** Maternal deaths reported in coroners’ PFDs published in England and Wales between July 2013 and August 2023 compared with deaths reported by MBRRACE in the UK between 2014 and 2021.

| Average over 3 years* | Deaths reported by MBRRACE in the UK | Deaths reported in coroners' PFDs in England and Wales | Percentage of MBRRACE-UK deaths written into a PFD (%) |
| --- | --- | --- | --- |
| 2013-15 | 528 | 9 | 1.7 |
| 2014-16 | 511 | 9 | 1.8 |
| 2015-17 | 522 | 13 | 2.5 |
| 2016-18 | 522 | 8 | 1.5 |
| 2017-19 | 475 | 10 | 2.1 |
| 2018-20 | 518 | 5 | 1.0 |
| 2019-21 | 552 | 6 | 1.1 |
| 2020-22 | NA | 6 | NA |
| 2021-23 | NA | 8 | NA |
| **Total 2013-21** | **2774** | **29** | **1.1%** |

\*A three year time period is used to align with MBRRACE reporting (Table 2.1 and 2.5, MBRRACE-UK Confidential Enquiries into Maternal Deaths 2013-21 reports)<sup>2</sup>

### Demographics

The median age of death was 33.5 years (IQR 28.5-36.0 years, n=20). The median years of life lost was 54 years per death (n=20, total=1051 years). One death occurred in a person aged <17 years, and eight (27.5%) in those aged >35 years.

Deaths most often occurred in hospitals (75.9%) followed by other community settings outside of the home (10.3%), and one in the home (3.4%). The location of death was not reported in three cases (10.3%).

Deaths occurred in 23 coroners’ areas out of a total of 82 areas in England in Wales, most frequently occurring in London Inner North (17.2%) (Supplementary Table S1).

The Chief Coroner’s Office (CCO) categorise PFDs into one or more types of deaths of which the most (78%) common was ‘Hospital death’ (Supplementary Table S2).

### Causes of deaths

The most frequent causes of death (Figure 1) reported by the coroner were haemorrhage (27.5%, n=8), followed by early pregnancy deaths (20.6%, n=6, which included complications of ectopic pregnancies and terminations), and suicide (20.6%; n=6). Further details of causes of death and ICD-11 coding can be found in Supplementary Table S2.

**Figure 1.**
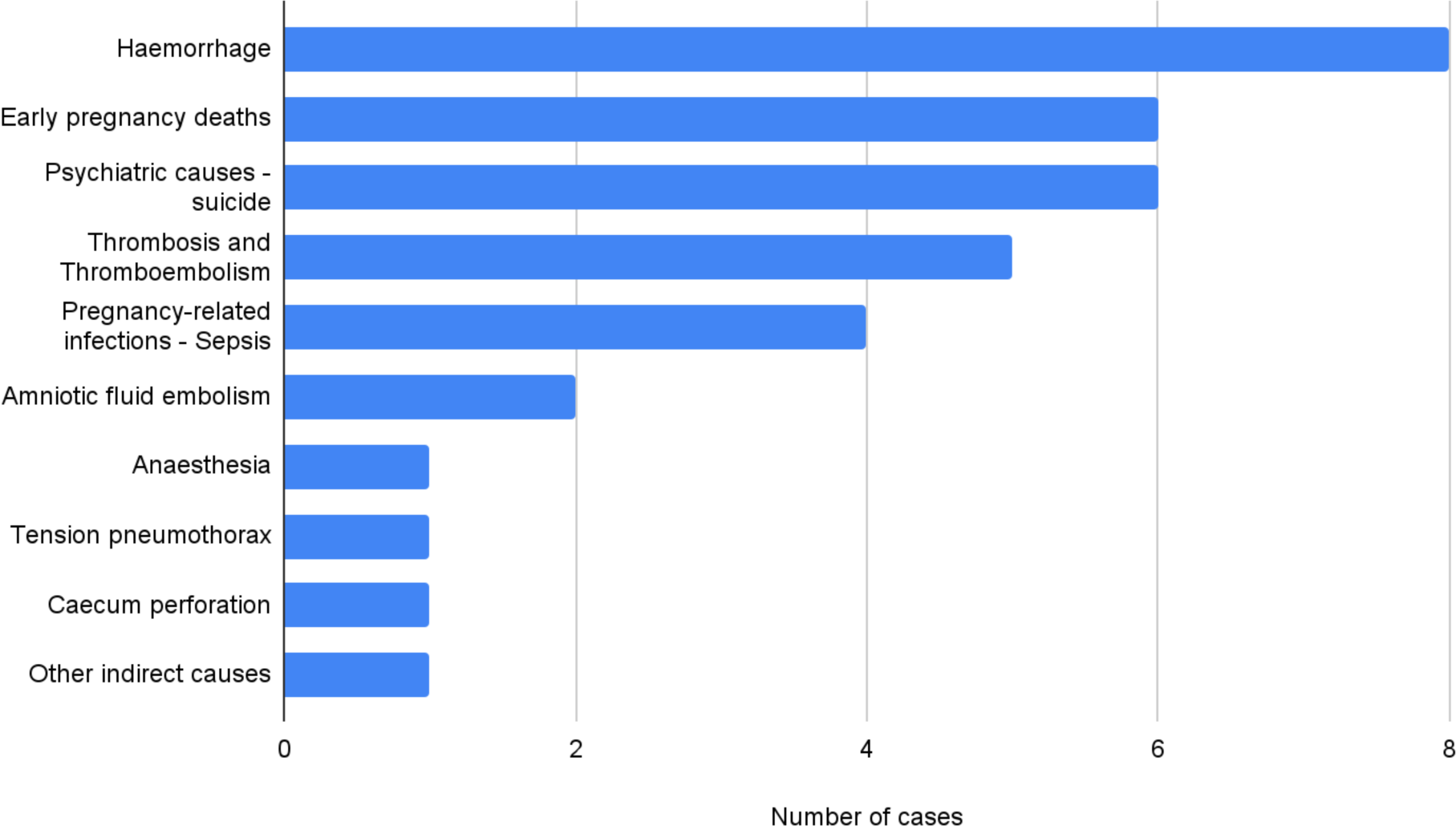
Causes of deaths reported by coroners in PFDs involving maternity deaths in England and Wales between July 2013 and August 2023. Causes of death from the 2019-2021 MBRRACE report were used to categorise deaths. Note: one case had >1 cause mentioned.

44.8% (n=13) cases reported none of the 15 factors associated with high risk pregnancies. 10.3% (n=3) of women had previous psychiatric history, and this was related to their cause of death. Other risk factors, including previous clotting disease and multiple pregnancies only occurred in a small number of individual (n=1) cases.

The majority (55.2%, n=16) of deaths occurred postpartum, including psychiatric causes and those occurring after abortion, or surgery for ectopic pregnancy. Antenatal deaths occurred in 24.1% (n=7) of women and intrapartum deaths (occurring during or within 24 hours of labour) occurred in 20.6% (n=6) of cases.

In 9 PFDs (31.0%) it was explicitly stated that pregnancy went on to lead to a live birth. Four (13.8%) of deaths involved ectopic pregnancies, three (10.3%) involved terminations of pregnancy, and one (3.4%) involved a miscarriage (<24 weeks). Two (6.9%) cases involved antenatal deaths of the mother in which the child was presumed to have died. In 10 cases, the outcome for the child was not mentioned.

### Coroner’s concerns

121 concerns were raised by coroners. The most common concern was regarding providing appropriate treatment (48.2%, n=14), the failure to escalate (37.9%, n=11), recognition of risk factors (31.0%, n=9), and lack of training (31.0%, n=9) (Supplementary Table S3). Specific lessons are discussed below in the next section.

### Responses to PFDs

PFDs were sent to 53 organisations. These organisations included NHS trusts (n=19) and professional bodies such as the General Medical Council or Medical Royal Colleges (n=13). Only 37.7% of PFDs received a response from the organisation to which they were sent. When organisations did respond, 8.00% (16 out of 20) of reported new changes including publishing new local policies, increasing multidisciplinary training in obstetric scenarios, or committing to increasing staffing levels (Supplementary Table S4).

The relation of types of concerns to whether change was initiated can be found in Figure 2. Specific actions taken in response to concerns can be found in Supplementary Table S1 and “Specific Lessons” above.

**Figure 2.**
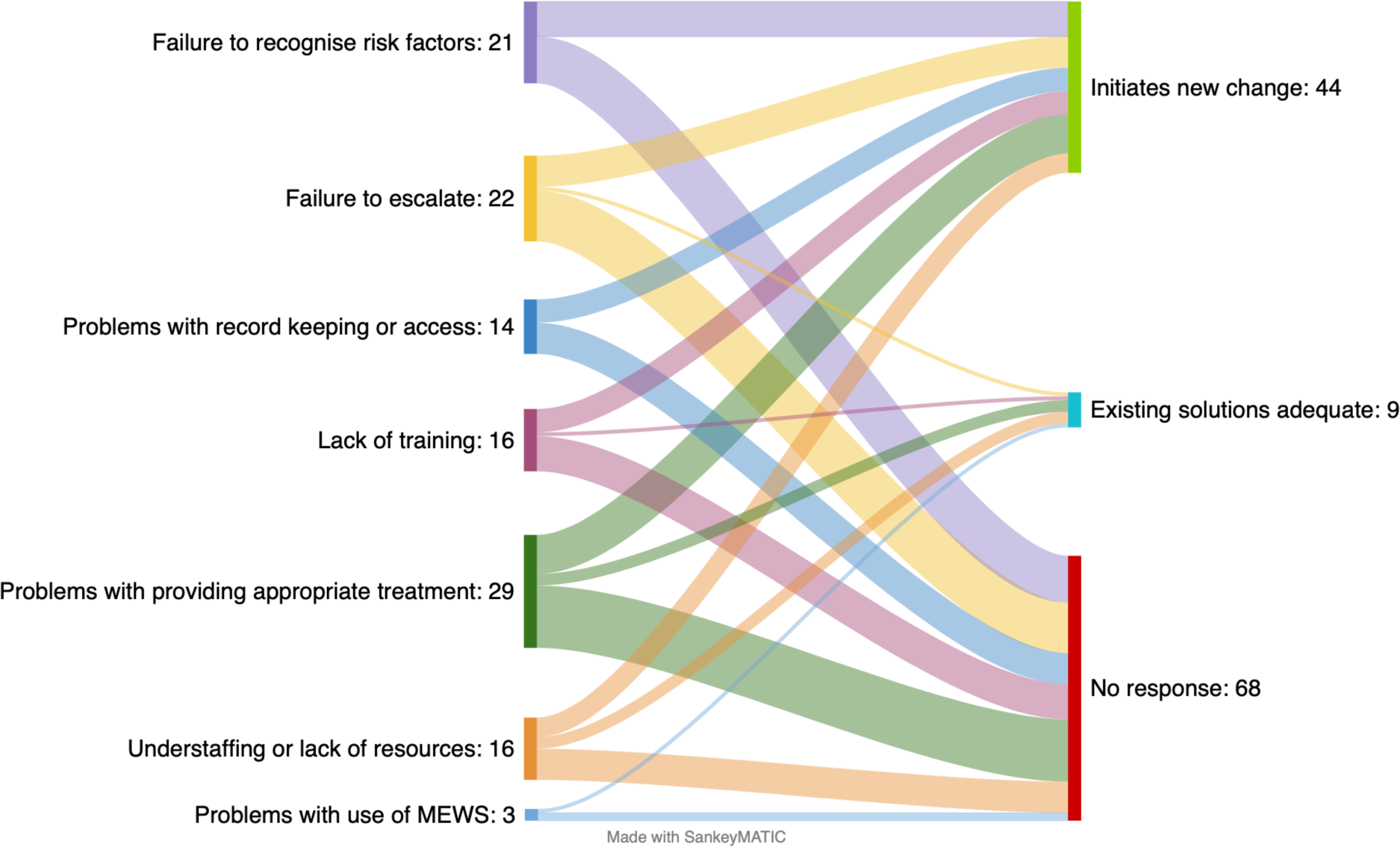
Concerns raised by coroners (left) in Prevention of Future Deaths reports involving maternity deaths in England and Wales published between July 2013 and August 2023 and the actions reported by organisations in their responses (right).

## SPECIFIC LESSONS

### Gaps in national guidance

PFDs highlighted gaps in national maternal care guidance. One case (2017-0005) concerned poor follow-up for discoloration found on amniocentesis, for which there was no existing guideline. The patient later presented with chorioamnionitis and died. The coroner’s suggestion to send such samples for immediate microbiological analysis was acknowledged by the Royal College of Obstetricians and Gynaecologists’ (RCOG) response with updated guidelines^21^.

Another patient (2021-0371) died due to sepsis from a retained fetus following feticide. The coroner highlighted the lack of guidance surrounding treatment of such infections, in particular whether antibiotics suffice, or if obstetric intervention is required. The hospital trust identified a similar death in the region and changed local guidelines. However, in lieu of national guidelines no specific treatment recommendations could be made.

A 2016 PFD detailed the death of a patient from bowel obstruction following previous bariatric surgery (2016-0213), where surgical causes of her symptoms were not adequately considered during pregnancy nor was a specific obstetric plan made. It was noted that there are no national guidelines for obstetric planning for pregnant women with previous bariatric surgery, a patient subset that is increasing in number. No published response from the RCOG, General Medical Council or Care Quality Commission was available so there were no responses to the concerns raised in this PFD.

### National protocols not followed

In a 2020 case (2020-0162), misoprostol administration to induce labour following intrauterine fetal death led to uterine rupture and maternal death. The hospital in question updated its misoprostol dose to match national guidelines and mandated medical reviews before misoprostol administration to multiparous mothers.

Four cases highlighted deficiencies in major obstetric haemorrhage protocols. In two cases (2015-0288; 2023-0095), local guidelines did not reflect national recommendations and were updated accordingly. A specific problem was blood products not being available near the maternity unit. In the other two cases (2022-0228; 2017-0020), local guidelines were not followed, leading to delays in diagnosis and treatment.

One death (2021-0418) resulted from a ruptured ectopic pregnancy where thrombolysis was administered for suspected pulmonary embolism (PE). While MBRRACE guidelines recommend a FAST scan in all women of child-bearing age suspected of a PE, this had not been integrated into local trust policy.

Another PFD (2015-0414) detailed failure to prescribe adequate doses of clexane for a mechanical valve contributing to development of fatal thrombosis. The report expressed concern from doctors that this was a recurring issue. There was no published response from the hospital trust or National Institute for Health and Care Excellence (NICE), so it is unclear if action was taken.

### Communication issues

In case (2015-0413), the patient attended the emergency departmentseven days postpartum, but obstetricians were not involved in her care, and information sharing between general practitioners, out of hours care and the obstetric department was poor. In another case (2019-0281), the ambulance crew did not communicate that the patient was pregnant to ED staff. In a third case (2019-0027), the obstetric department was not forewarned about a pregnant patient’s arrival to ED, leading to delays in care. Two actions were reported by trusts in their responses, including the addition of a prompt to record pregnancy status on call sheets and the development of a set of criteria to determine if an obstetric call needs to be initiated prior to ambulance arrival.

Poor communication between teams was highlighted in the psychiatric care of pregnant women. Two PFDs reported failures to coordinate multidisciplinary care for pregnant patients between their medical and psychiatric teams (2015-0418, 2017-0055). Delays in accessing psychiatric care were found to directly contribute to the death of another patient (2022-0303).

### Lack of resources or staff-cover

Concerns have also been raised regarding staffing and resource allocation. In one case, there was no formal method of getting assistance when the first consultant called could not attend an emergency (2019-0453). Another challenge appears to be regions with poor availability of perinatal health clinics. This was the case in one PFD (2014-0239), in which the mother was not able to access community care that met both her and her son’s needs. The report claimed that in the region mentioned, 50% referrals to the nearest Mother and Baby Perinatal Mental health in-patient Unit are declined due to distance.

## DISCUSSION

### Main Findings

We identified 29 maternal deaths reported by coroners in PFDs. There was a broad range of causes of death, occurring mostly in the postpartum period in hospitals. Coroners raised significant concerns spanning all stages of pregnancy, but only 38% of PFDs received a response. PFDs frequently highlighted gaps in national guidance, lack of consistency in local guidelines, and problems with communication.

### Strengths and Limitations

This study uses reproducible methods from previous research of PFDs involving other types of deaths^12^. However, to the best of our knowledge, it is the first published study of PFDs involving maternal deaths reported by coroners’ in PFDs.

We compared the data from coroners with the MBRRACE-UK initiative – an established national data source on maternal deaths in the UK^2^. Our findings illustrate the ability of coroners’ reports to provide unique case-level insights into issues in care, systems, and processes, which complements larger scale epidemiology research such as MBRRACE-UK.

Limitations of the PFD data are well-established^22^, including inter-coroner and inter-regional variability in the publication of reports and the information reported in them. In the maternal setting, specifically, the type of deaths identified by coroners may be biased by those sent for autopsy. This has resulted in a small sample size of maternal deaths which may not be representative of maternal deaths as a whole in the UK.

PFDs also do not consistently report established factors contributing to maternal health inequality, including ethnicity, socio-economic status, and previous parity. It is difficult to conclude that risk factors were not present in a PFD case, as there is no requirement for coroners to consistently report them.

### Interpretation

Maternal deaths continue to be major global health issue^23^. As emphasised in a recent call to action by the International Network of Obstetric Survey Systems (INOSS), one of the key steps to address stagnating maternal mortality rates is learning from case-based analyses of maternal deaths^24^. National analyses in the USA ^25^ and China ^26^ suggest that over 80% of maternal deaths may be preventable.

Coroners in the UK are able to report maternal deaths to MBRRACE-UK ^2^, which collates these deaths for the confidential enquiry into maternal death and morbidity. Separately, coroners have a duty to write PFDs, but we found that only around 1% of maternal deaths in the UK reported by MBRRACE were written into a PFD. This is an underestimate of the true number of maternal deaths where action ought to be taken by organisations.

Coroner reports can provide unique insights into the systems and processes that can go wrong and lead to preventable deaths. An analysis of coroner reports of maternal deaths in Ontario, Canada^27^ found that (when physical injury was excluded), the two most common causes of death were haemorrhage and suicide, both of which were in the top three causes of death seen in our study^27^.

Many of the concerns raised in PFDs were reflected in the most recent MBRRACE-UK^2^ report, including failures in providing appropriate treatment, recognising risk factors, and communication issues, which is an important aspect of maternal care highlighted in the wider literature^28^. Similar concerns found in our analysis and MBRRACE-UK included a need to have accessible electronic records, better shared management care across multidisciplinary teams, and identification of care coordinators when multiple teams are involved. A thematic analysis of maternal deaths in London, the region with the highest number of maternity-related PFDs, conducted by the London Maternity Network’s Maternal Morbidity and Mortality Working Group, had similar messages including improved adherence to protocols and improved access to care for women with complex medical needs^29^.

## CONCLUSION

PFDs are an under-recognised source of data for improving maternal care and reducing preventable deaths. Organisations that receive PFDs from coroners may be failing to take action as there is no mechanism to follow up on missing responses or ensure that reported actions are implemented. Using PFDs, we identified issues in the provision of care and gaps in national guidance and policy. We have created a reproducible method for collecting and analysing reports (https://preventabledeathstracker.net/), so that PFDs can be more widely used as a learning tool to prevent future deaths. To improve access to such reports, the Chief Coroner’s Office should consider updating their categorisation of deaths and include ‘Maternal deaths’ as an official classification for PFDs.

## Acknowledgements

None.

## Contribution to authorship

GCR established the methodology used to perform case series’ of PFDs (https://preventabledeathstracker.net/). JJ and DL conceived of the study and wrote the protocol. FD wrote the code used to download the pdf documents and performed the keyword screening using the OCCRP Aleph tool. JJ and DL performed data extraction and analysis. All authors interpreted the study findings and contributed to writing and reviewing the manuscript. All authors accept responsibility for the paper as published.

## Details of Ethics Approval

This study uses publicly available information, for which ethics committee approval is not required. Both the General Data Protection Regulation (GDPR) and the Data Protection Act (2018) no longer apply to identifiable data that relate to a person once they have died(30).

## Data availability

Protocols and study materials used for data synthesis are openly available on the Open Science Framework (https://osf.io/7h4j6/). Demographic information from all PFD cases is openly available on the Preventable Deaths Tracker (https://preventabledeathstracker.net). The code used to download all the PFD pdf documents is openly available (https://github.com/francescodernie/coroner_PFDs).

