## Supplementary Information for "Preventable maternal deaths in England and Wales, 2013-2023: a systematic case series of coroners’ reports"

**Supplementary Table S1.** Classification of sepsis-related Prevention of Future Death reports (PFDs) according to coroner area described in the report.

| **Coroner Area** | **Number of PFDs** |
| --- | --- |
| Avon | 1 |
| Bedfordshire & Luton | 1 |
| Berkshire | 1 |
| Birmingham and Solihull | 1 |
| Blackpool & Fylde | 1 |
| Derby and Derbyshire | 2 |
| Gloucestershire | 1 |
| Gwent | 1 |
| Liverpool and Wirral | 1 |
| London (East) | 2 |
| London (North) | 5 |
| London (West) | 1 |
| Lincolnshire | 1 |
| Manchester (North) | 1 |
| Manchester (South) | 1 |
| Norfolk | 1 |
| North West Kent | 1 |
| Nottinghamshire | 1 |
| South Yorkshire (East) | 1 |
| Surrey | 1 |
| West Sussex | 1 |
| Worcestershire | 1 |

**Supplementary Table S2. Summary of the 29 Coroners’ Prevention of Future Deaths (PFDs) reports involving maternal deaths in England and Wales between July 2013 and August 2023 by case number, cause of death*, and response if present**

| **Case** | **Medical Causes of Death reported by Coroner** | **Asssigned MBRRACE cause of death** | **ICD-11 codes** | **Chief Coroner’s Office categoisation** | **Summary of Concerns** | **Changes made if Response Present** |
| --- | --- | --- | --- | --- | --- | --- |
| 2016-0213 | 1a: ARDS and aspiration pneumonitis, small bowel infarction 1b: gut obstruction (operation) 1c: previous bariatric surgery | Other indirect | CB00, DD30 | Hospital Death (Clinical Procedures and medical management) related deaths | 1) Lack of obstetric consultant supervision in care  2) Poor documentation of observations and fluid chart  3) No specific obstetric plans for patient despite having undergone bariatric surgery, and lack of RCOG guidelines in this matter  4) Surgical causes of sudden abdominal pain not considered in pregnant patient with surgical history | Ashford and St Peter’s Hospital: Introduces a change in consultant working practices. Has created a new divisonal guidlines on pregnant women who have previously had bariatric surgery. Introduced and reviewed an audit on the relevant ward, and references current and emerging royal college guidlines on mothers post-surgery. |
| 2019-0281 | 1a: hypovolaemic shock due to massive intra abdominal bleeding (emergency laporotomy) 1b: rupture of the abnormal gravid uterus 2: monochorionic diamniotic pregnancy (18/40) | Haemorrhage | JA40 | Hospital Death (Clinical Procedures and medical management) related deaths | 1) Pre-alert call from ambulance did not communicate patient was pregnant  2) Even after arrival at hospital, it took 16 minutes post arrival for the pregnancy to be recognised and the obstetric team to be called.  3) Without obstetrics, the ED on the potential for pulmonary embolism and alteplase was given. Only later was a scan conducted and free fluid noted. By the time of laparotomy it was too late to save the patient. | London Ambulance Service: Extensively reviews the ambulance communication in the case. Introduces new training sessions including ambulance staff.  Whittington Health NHS trust: Modifies pro-forma for taking ambulance calls, and criteria for involvement of obstetricians. Has new simulation training exercises for obstetric patients. |
| 2017-0096 | 1a: shock and haemorrhage due to 1h: perforated gastric ulcer | Haemorrhage | MG40 | Community health care and emergency services related deaths | [not related to death directly]  1) Circumstances in which methadone was prescribed - over phone without medical review and without details by which patient was taking pain relief | No Responses |
| 2023-0174 | Pulmonary embolism due to DVT, risk of which was increased by her Klippel–Trenaunay syndrome | Thrombosis | BB00 | Hospital Death (Clinical Procedures and medical management) related deaths ; Emergency services related deaths (2019 onwards) | 1) Lack of communication between teams in Sheffield and Chesterfield about patient's anticoagulation medication, resulting in her not receiving it after moving  2) Lack of awareness about patient's syndrome (Klippel–Trenaunay syndrome) - however it was not documented in her notes or its implications on pregnancy considered | No Responses |
| 2015-0414 | Multi organ failure due to acute thrombosis of the mechanical mitral valve in the first trimester of pregnancy due to emergency cardiac surgery for acute decompensation due to rheumatic mitral valve disease after previous pregnancy | Thrombosis | BB6Y | Hospital Death (Clinical Procedures and medical management) related deaths | 1) Failures on part of cardiology team to adequately investigate complications of patient's mechanical valve ie. delays in performing echocardiogram; consultant review was not sought  2) Failure to prescribe adequate doses of clexane contributed to development of fatal thrombosis - noted by consultants that this may be recurring problem in those switched over from warfarin  3) Lack of understanding amongst staff of increased thrombosis risk for pregnant women with mechanical valves | No Responses |
| 2014-0373 | 1a: raised intracranial pressure 1b: cerebral haemorrhage | Haemorrhage | 8D60 | Hospital Death (Clinical Procedures and medical management) related deaths | 1) CT scan was misinterpreted - they were reviewed by a radiologist not a neuro-radiologist which guidelines at the hospital specify should have happened | No Responses |
| 2017-0005 | 1a Escherichia coli sepsis  1b choriamnionitis  1c second trimester pregnancy. | Pregnancy-related sepsis | 1G40 | Hospital Death (Clinical Procedures and medical management) related deaths | 1) Amniocentesis had found discoloured blood but this was not followed up in absence of Infectious symptoms. Patient later presented with chorioamnionitis and died due to complications. Given the rarity of such non blood stained discolouration, it may be a wise precaution in this situation always to send a sample for immediate microbiological analysis, and quickly to follow up the result. | British Maternal and Fetal Medicine Society: Agrees with need to send discloured amniotic fluid. Suggests that current guidelines on amniocentesis are dated. |
| 2023-0095 | Placental haemorrhage and amniotic fluid embolism due to diagnosed placenta praevia | Haemorrhage, amniotic fluid embolism | MG27, JB42 | Hospital Death (Clinical Procedures and medical management) related deaths | 1) Obstetric plan not recorded in notes, poor note keeping overall  2) Trust had not adopted guidance for delivery by caesarean section between weeks 36-37 in Chloe’s circumstances  3) Delay in provision of blood products due to lack of robust major obsetetric haemorrhage protocol and blood products for urgent use not being kept near maternity unit  4) Delay in acquiring anaesthetist after caesarean had been decided on  5) Lack of key equipment on maternity ward including warming equipment for women during surgery, point of care resting for anticoagulation and blood storage fridge | The University Hospitals of Derby and Burton: Commissioned a broad audit into the governance of the trust and maternity services. Investing in additional maternity staffing and funding. |
| 2017-0020 | 1a: cardiopulmonary arrest 1b: problems relating to general anaesthesia 1c: recent third trimester delivery, sepsis, and acute kidney injury | Sepsis, Anaesthsia | MC82 | Hospital Death (Clinical Procedures and medical management) related deaths | 1) No actions taken to check and ensure that no part of placenta remains following a C Section delivery  2) Protocol for management of post partum haemorrhage was not followed by medical staff  3) Delays in acquiring anaesthetist/intensivist  4) Poor note taking | Maidstone and Tunbridge Wells NHS Trust: Makes extensive reference to protocols and trainings already in place in the trust. Introduces rota changes for caesarian section anaesthetic cover. |
| 2016-0117 | Perforated Caecum (Ogilvie’s syndrome) - rare complication of Caesarian Section | Direct - other | ME24 | Hospital Death (Clinical Procedures and medical management) related deaths | 1) Modified obstetric early warning score tool was not used appropriately to identify Ms Ganesh-Ram’s sepsis  2) Delays in obtaining CT scan  3) No surgical consult sought for 2 days after symptoms despite CT showing large fluid volume in peritoneum  4) Signs of perforated caecum (raised pulse, abdominal pain and lack of urine output) were not adequately escalated | No Responses |
| 2015-0288 | Amniotic fluid embolism | Amniotic embolism | JB42 | Hospital Death (Clinical Procedures and medical management) related deaths | 1) Major obstetric haemorrhage protocol did not meet national guidelines  2) Medical staff and porters were unaware of Health Board protocols | No Responses |
| 2019-0453 | Disseminated Intravascular Coagulation after PPH | Haemorrhage, thrombosis | 3B20, JA43 | Hospital Death (Clinical Procedures and medical management) related deaths | 1) Lack of confidence/ability on part of on call consultant to perform emergency total abdominal hysterectomy without another consultant present - this was not the practice at other hospitals to have two consultants present. There was also a lack of leadership of patient's care including reluctance on part of on-call consultant to consider anyting other than conservative measures until another obstetric consultant was present.  2) There was no formal method of getting assistance when first consultant called could not attend  3) Lack of professional curiosity about the cause of DIC as haemorrhage was not enough to cause this | No Responses |
| 2023-0014 | 1a. Acute Bronchopneumonia  1b. Global Cerebral Hypoxia  1c. In Hospital Cardiac Arrest following Third Trimester Lower Segment Caesarean Section, Significant Post-Partum Haemorrhage and Perioperative Bilateral Tension  Pneumothoraces | Haemorrhage, direct-other | CA40, MC82 | Hospital Death (Clinical Procedures and medical management) related deaths | 1) Tension pneumothorax was not considered as cause for Pulseless Electrical Activity Cardiac Arrest, despite it being the only cause resulting in sudden inability to ventilate  2) Delay in the recognition of surgical emphysema despite clinical signs  3) Despite patient being intubated and ventilated at time of event, there were no steps taken by anaesthetic department to investigate potential iatrogenic or other anaesthetic related causes, take the anaesthetic equipment out of service to investigate possible faults or download data from the anaesthetic machine to establish how patient suddenly developed bilateral tension pneumothoraces  4) Lack of a proper and robust system in place to trigger an investigation into all the circumstances of the death of a 17-year-old patient | Royal College of Anaesthetists: Reviewed and change guidance in investigative processes for catastrophic anaesthetic events |
| 2022-0228 | Complications of uterine inversion leading to cardiac arrest and major uterine haemorrhage | Haemorrhage | MC82, JA42 | Hospital Death (Clinical Procedures and medical management) related deaths | 1) Insufficient support for newly appointed Obstetric Consultants  2) Concerns regarding the conduct of the Trust Serious Incident Investigation  3) Management of the uterine haemorrhage was not undertaken as per guidelines, with delay in administration of uterotonic drugs or identification of bleeding source | The Chief Executive, Doncaster and Bassetlaw Teaching Hospitals NHS Foundation Trust: Changes to involve family more directly with mortality investigations. Introduced a new training courses delivered by the Healthcare safety investigations branch. Very detailed review of situations regarding investigation into the case. New protocols for consultants supporting other consultant colleagues. |
| 2015-0413 | Ruptured splenic artery aneurysm | Haemorrhage | BD51 | Hospital Death (Clinical Procedures and medical management) related deaths | 1) Lack of escalation when patient developed chest pain after giving birth  2) No process to ensure sharing information between midwives, out of hours, GP and obstetric department  3) When patient attended ED as a post natal patient (7 days post partum), obstetric department were not involved in her care | No Responses |
| 2018-0302 | Pulmonary embolism | Thrombosis | BB00 | Hospital Death (Clinical Procedures and medical management) related deaths; Community health care and emergency services related deaths | 1) Signs of venous thromboembolism not recognised by GP  2) Poor record keeping at GP practice  3) Insufficient medical registrar cover at hospital during nighttime | No Responses |
| 2019-0027 | 1a: haemoperitoneum 1b: ruptured ectopic pregnancy of the left fallopian tube | Early pregnancy causes | JA40 | Hospital Death (Clinical Procedures and medical management) related deaths; Emergency services related deaths (2019 onwards) | 1) Delay in ambulance arrival time  2) Despite pre-alert calls by parademics to serious nature of patient's condition, clinicians were not ready for her arrival. The pre-alert calls were also recorded but not dated or signed.  3) O&G department was not forewardned about pregnant patient's arrival to ED | No Responses |
| 2021-0418 | Ruptured ectopic pregnancy | Early pregnancy causes | JA01 | Hospital Death (Clinical Procedures and medical management) related deaths | 1) FAST scan did not take place before diagnosis of pulmonary embolism made - lead to administration of alteplase in patient with intra-abdominal bleeding  2) MBRRACE 2019 guidelines that "Women of reproductive age, presenting to the ED collapsed, in whom a pulmonary embolism is suspected, should have a Focussed Assessment with Sonography in Trauma (FAST) scan to exclude intra-abdominal bleeding from a ruptured ectopic pregnancy especially in the presence of anaemia" is now adopted into local Trust policy but has not been incorporated into National Resuscitation Council UK, Obstetric Cardiac Arrest guidance  3) Concern that obstetricians do not receive training to identify intra-abdominal bleeding | Royal College of Obstetrics & Gynaecology: Makes reference to current RCOG guidelines and practices for scanning mothers during early pregnancy, but does not suggest changes to practices.  Director of Clinical & Service Development, Resuscitation Council UK: Introduces a number of alterations to resuscitation guidelines and training materials relevant to pregnancy, informed by MBRRACE |
| 2015-0126 | Ruptured ectopic pregnancy | Early pregnancy causes | JA01 | Hospital Death (Clinical Procedures and medical management) related deaths | 1) Delay in ambulance arrival time; potential for systems improvements, such as automated recategorisation, clinical re­triaging and feedback to call­handlers regarding current time­frames  2) Guidance and training for ambulance service does not endorse testing for pregnancy on scene for all women of childbearing age with abdominal pain  3) Insufficient knowledge of alternative extraction techniques by ambulance crew  4) Potential for an ‘early warning score’ system, which is specifically validated for pre­hospital use  5) Concerns regarding governing and internal investigations by ambulance system | NHS England: Discussed multiple reviews commissioned of different services involved with the case.  College of Paramedics: Made commitment that concerns will be brought up in meetings with other collaborative bodies, advice given and relevant new guidance initiated.  London Ambulance Service NHS Trust (LAS): Detailed review of each concern, explaining decisions regarding ambulance funding, pre-hospital pregnancy testing, patient evacuation from the scence, and systems-level communications between call handlers, clinical governance, and early warning score use. |
| 2017-0129 | Bilateral pulmonary thromboemboli | Thrombosis | BB00 | Hospital Death (Clinical Procedures and medical management) related deaths | 1) Failure of doctors to diagnose pulmonary embolism  2) Failure of nurses notes to be provided to doctors  3) Deficiencies in operating of NEWS score and training of clinicians using it | No Responses |
| 2020-0162 | Uterine rupture caused by the admistration of misoprostol prescribed to induce labour following diagnosis of intrauterine death | Early pregnancy causes | JB0A | Hospital Death (Clinical Procedures and medical management) related deaths | 1) The misoprostol was administered at doses in excess of the Royal College of Obstetricians and Gynaecologists national guidelines  2) Failure of clinician to attend to patient despite midwife reporting abnormal observations  3) Lack of computer in delivery suite meant midwife had to leave room to update notes or record observations  4) Insufficient dissemination of learnings nationally that there is increased increased risk of uterine rupture in a multi gravida mother | Royal Free Hospital Trust: Case presented locally at safety meeting, learning disseminated locally. Protocols to ensure computer present in each delivery suite. |
| 2021-0371 | 1a: multiorgan failure 1b: sepsis 1c: feticide for trisomy 21 | Sepsis, early pregnancy | 1G40 | Hospital Death (Clinical Procedures and medical management) related deaths | 1) Lack of Informed consent and maternal choice regarding mode of delivery, poor documentation of discussions regarding mode of delivery, maternal wishes and risk/benefits of differing management plans.  2) Concern that infection risk of retained foetus following feticide is not being given weight in clinical decisions. There does not appear to be any local or national guidance considering whether infection is controlled by antibiotics alone, whether swifter methods of foetal delivery (ie. caesarean section) are required or when multidisciplinary advice is warrented | Worcestershire Acute Hospitals NHS Trust: Number of new protocols within obstetric department relevant to communication between staff and recording keeping. |
| 2022-0144 | 1a Group A Streptococus Sepsis following Medical Termination of Pregnancy | SEpsis, early pregnancy | 1G40 | Hospital Death (Clinical Procedures and medical management) related deaths, Community health care, Other related deaths | 1) Inadequate training of doctors and other medical professionals re the risk of sepsis following Early Medical Terminations  2) Poor note taking at GP surgery meant pharmacist who saw patient was not aware she had recently undergone early medical abortion  3) Sepsis was not recognised or treated by the GP surgery, emergency department or Acute Medical Unit and upon Sarah’s arrival at hospital, the sepsis pathway was not followed | Department of Health & Social Care: Makes reference to pre-existing and upcoming sepsis early diagnosis tools, as well as GMC guidelines on doctor education. |
| 2022-0303 | (Not reported by coroner) Presumed suicide by jumping | Suicide | PD3Z | Suicide (From 2015) | 1) The British Pregnancy Advisory Service (BPAS) is a charity whose services are often commissioned by the NHS. As a charity, BPAS does not have direct access to NHS perinatal psychiatrists. Referrals would have to be made either via the patient’s GP or via an unwieldy safeguarding concern (as happened in this case). Referrals via the GP are not possible where the patient does not wish their identity to be revealed. | No Responses |
| 2017-0163 | 1a: hypoxic ischaemic brain injury 1b: cardiac arrest 1c: trauma to the liver | Suicide | PD3Z | Hospital Death (Clinical Procedures and medical management) related deaths | 1) Poor identification of physically unwell patient in an acute mental health setting prevented an escalation in care. MEWS score of 1 was not acted upon. Very few nurse consultants in physical healthcare working in mental health settings. | NHS England: References a range of pre-existing programmes within NHS England to support the physical and mental health of mothers |
| 2015-0418 | 1a: multiple injuries | Suicide | PD3Z | Hospital Death (Clinical Procedures and medical management) related deaths | 1) No multidisciplinary team meeting (ie. with GP, midwife, obstetrician, psychiatrist, care coordinator, social services) when patient with high risk mental health condition became pregnant  2) There was no appropriate care plan formulated and circulated to those involved in patient treatment  3) Concerns when the patient stopped taking her Risperidone during pregnany were not followed with any meeting with psychiatric care  4) When the patient deteriorated and suffered a relapse there was a failure to diagnose and manage this | Avon and Wiltshire Mental Health NHS Trust: Prepares a new training exercise based on the case, and reviewed perinatal pathways within the trust. |
| 2013-0309 | 1a: multiple injuries | Suicide | PD3Z | Mental Health related death | 1) Concerns are surrounding infrastructure and security at her residence which allowed her to commit suicide | No Responses |
| 2017-0055 | 1a: severe anoxic brain injury | Suicide | PD3Z | Hospital Death (Clinical Procedures and medical management) related deaths, Mental Health related deaths, Suicide (from 2015) | 1) Despite patient and her family making it clear to staff that they wanted her medication changed, no medication review took place for 4 days she was an inpatient before her death  2) Concerns regarding Rache's risk of self harm/suicide did not generate full risk assessment to be conducted  3) When reviews conducted, these were not reviewed by MDT as per policy  4) Lack of clarity around different levels of observations needed within Trust policy  5) Overreliance of patient being inpatient as a protective factor | No Responses |
| 2014-0239 | Ia Hypoxic Brain Injury  Ib Hanging | Suicide | PD3Z | Mental Health related death | 1) Lack of Specialist Community Perinatal Mental Health Service in region (Wirral)  2) No Mother and Baby Perinatal Mental health in–patient Unit in the Liverpool City Region serving the needs of Lancashire, Merseyside and East Cheshire. 50% of referrals from this area to the Manchester Unit decline because it is too far from family and support networks but more relevantly from older sibling children who remain in the family home. | Wirral Clinical Commissioning Group: Investigation into current maternity provision. Additional recruitment for perinatal midwife, and input increased from consultant psychiatrist into maternity.  Department of Health: Details the organisation of local maternal mental health services. |

**Supplementary Table S3. Classification of coroner concerns**

| **Concern Type** | Number of PFDs | Percentage |
| --- | --- | --- |
| Problems with providing appropriate tratment | 15 | 51.7% |
| Failure to escalate | 11 | 37.9% |
| Failure to recognise risk factors/comorbidities - including recognition of high risk pregnancy | 9 | 31.0% |
| Lack of training | 9 | 31.0% |
| Problems with accessing records, or keeping accurate records | 8 | 27.6% |
| Understaffing, lack of resources | 6 | 20.7% |
| Problems with use of Maternal Early Warning Score | 2 | 6.9% |
| Poor discharge planning or lack of follow up | 0 | 0.0% |

**Supplementary Table S4.** Classification of types of response from organisations who responded to coroner concerns raised in maternal death-related PFDs

| **Responses** | **Number of responses** |
| --- | --- |
| Responds, Acknowledges concern and initiates new change to address concern | 16 |
| Responds, Acknowledges concern but says pre-existing systems/solutions adequate | 4 |
| Responds but does not acknowledge/agree with concern | 0 |
| No response | 33 |
| **Total** | **53** |

**Supplementary Table S5.** Organisations and individual who received maternal death-related PFDs, and their response rates according to the statutory requirement of 56 days.

| **Organisation Type** | **Total PFDs Received (n)** | **Responses Received (n)** | **Response rate (% of PFDs responded to)** | **On time responses** | **Late responses** | **Undated** | **Overdue responses** |
| --- | --- | --- | --- | --- | --- | --- | --- |
| NHS CCG | 2 | 1 | 50.0% | 1 | 0 | 0 | 1 |
| NHS England | 7 | 2 | 28.6% | 0 | 2 | 0 | 5 |
| NHS Ambulance Services | 2 | 2 | 100.0% | 2 | 0 | 0 | 0 |
| NHS Trusts & Hospitals | 19 | 7 | 36.8% | 4 | 1 | 2 | 12 |
| NHS GPs | 2 | 0 | 0.0% | 0 | 0 | 0 | 2 |
| Welsh Health Boards | 1 | 0 | 0.0% | 0 | 0 | 0 | 1 |
| NHS Mental Health Trusts & Clinics | 1 | 1 | 100.0% | 1 | 0 | 0 | 0 |
| Government Bodies/Departments | 2 | 2 | 100.0% | 1 | 1 | 0 | 0 |
| Professional Bodies | 13 | 5 | 38.5% | 3 | 1 | 1 | 8 |
| Other ie. private companies, local councils | 4 | 0 | 0.0% | 0 | 0 | 0 | 4 |
| **Total** | **53** | **20** | 37.7% | **12** | **5** | **3** | **33** |

**Supplementary Table S6.** Classification of improvements made by organisations who responded to maternal death-related PFDs and initiated changes in response

| **Changes initiated** | **Number of responses*** |
| --- | --- |
| Improve patient information | 1 |
| Improved training / re-education | 8 |
| Initiation of audits or investigations | 9 |
| Improvements in communication / handover processes | 1 |
| Commitment to increase staffing levels | 5 |
| Improvement / implementation of new protocols, pathways or guidance documents | 12 |
| Changes to record-keeping / note-taking / medication monitoring | 3 |
| Investment / increase in resources / resource-reallocation (including IT systems) | 3 |
| **Total** | **42** |

*Note Responses may mention more than one type of change
